# The comparison of Post-Operative Complications pre Covid era versus during Covid-Era based on Clavien-Dindo-classification: A Systematic Review and Meta-Analysis

**DOI:** 10.1101/2022.02.25.22271519

**Authors:** Yeganeh Farsi, Fatemeh Shojaeian, Seyed Amir Ahmad Safavi-Naini, Mohammadjavad Honarvar, Benyamin Mohammadzadeh, Mohammad Javad Nasiri

**Author notes:** Theses authors are equally considered as the first author.

## Abstract

**Introduction:** Coronavirus Disease 2019 (COVID -19) pandemic challenged the healthcare system drastically, and it was concomitant with a remarkable decline in surgeries and modified routine care of patients worldwide. This systematic review and meta-analysis aimed to compare the surgical complications before COVID -19 (Pre-COVID) and after COVID -19 (post-COVID) appearance using the Clavien-Dindo classification (CDC).

**Methods:** between January 1, 2019, to November 3, 2021, we performed a comprehensive search in PubMed/Medline and Scopus for studies reporting the postoperative complications based on/transformable to CDC.

**Result:** From 909 screened articles, 34 studies were included for systematic review. Among included articles, 11 were eligible for meta-analysis. Nineteen thousand one hundred thirty-seven patients (pre-COVID: 3522, post-COVID: 15615) were included, mostly undergoing elective surgeries (86.32%). According to CDC classification, there were no significant change between pre-COVID and post-COVID for grade 1 (Odds ratio (OR) and 95% confidence interval (95-CI): 0.99, 0.60-1.63, p=0.96), grade 2 (OR and 95-CI: 0.65, 0.42-1.01, p = 0.055), grade 3 (OR and 95-CI: 0.86, 0.48-1.57, p=0.64), grade 4 (OR and 95-CI: 0.85, 0.46-1.57, p =0.60). However, the postoperative mortality was lower before the COVID -19 outbreak (OR and 95-CI: 0.51, 0.27-0.95, p= 0.035). The included studies for systematic review and meta-analysis had a low risk of bias and unsignificant publication bias.

**Conclusion:** Although delivering routine surgery was challenging, the postoperative complications during the pandemic remained identical to the pre-pandemic era. The stricter patient selection tending to choose more critical states and more advanced clinical stages of the operated patients may explain some extent of higher mortality during the pandemic. Adopting preventive strategies helped deliver surgeries during the outbreak of COVID -19 while limiting the capacity of operations and admissions.

## Introduction

Coronavirus Disease 2019 (COVID -19), which was declared as a pandemic in March 2020, has expanded fast worldwide since its emergence in late December 2019 and claiming more than 5.6 million lives (1–3). The pandemic was wreaking havoc on all aspects of healthcare systems by overwhelming them with contagious patients and a newly appeared disease (4,5). Sick patients with various signs and symptoms flooded hospitals and other medical institutions (6,7); hence, they were unable to provide a wide range of procedures and surgeries while still ensuring the health and safety of patients and staff. Accordingly, an international call to postpone surgeries seemed crucial for as many as possible (8,9).

The world and healthcare organizations have been gradually returning to normal as the virus has been contained. Since the surgical candidates were already facing prolonged delays, resuming surgeries was one of the earliest steps of this returning (10,11). As a result, different hospitals have recommenced surgeries one by one, and the number of invasive procedures is rising all over the world. By continuing the reduction of lockdown restrictions, as well as flattening the exponential curve of the virus spread and its related death, a long waiting list for different types of procedures would be expected (12,13). Evidently, the pandemic is ongoing with no end in sight, and the virus would continue to rule the healthcare systems. Considering some strategies appeared for patients’ safety and protection, there would be a trade-off between the risk of performing surgeries during the pandemic and further postponement or cancellation (14,15).

Taken together, CVID-19 seems to continue infecting individuals and running the healthcare systems and their policies; thus, there might be an urgent need to investigate whether resuming the surgeries was a drawback to the health condition of the patients and resulted in more surgical complications or not. Here, we are conceived to go into the complications of the patients who underwent surgery during the pandemic. Particularly, in this systematic review, we aimed to compare the surgical complications in various types of surgery, including urgent, emergent, and elective surgery, before and after COVID -19 appearance, using the Clavien-Dindo classification as a valuable, objective rating method for surgical complications (16).

## Methods

### Search Strategy

As we aimed to determine the outcomes of surgical procedures during the COVID era, we only considered publications between January 1, 2019 to November 3, 2021. The searched keywords included: COVID -19, SARS CoV 2, Coronavirus Disease 19, Surgery, operative therapy, invasive procedure, operative procedure, operation, perioperative procedure, intraoperative procedure, preoperative procedure, Clavien-Dindo, and postoperative complication. The study is performed based on the latest edition of PRISMA guidelines (17).

### Study Selection

The primary search results were organized by EndNote X8 (Thomson Reuters, New York, NY, USA) and duplicates removed. Two independent reviewers (FS and BM) evaluated the relevancy of the remaining articles based on the title and abstract. The remained full-text were assessed by the third reviewer (YF) and the data of eligible articles were extracted by two other reviewers (AS and MH). The eligibility criterions for systematic review study selection were defined as: 1) Reporting the post operation complications based on Clavien-Dindo-Classification explicitly or transformable to Clavien-Dindo-Classification. 2) Study population as adult patients with negative pre operative RT-PCR of SARS-CoV-2 or unprobeable cases of COVID -19. 3) Comparing the post operative complications before and during the COVID era was an additional inclusion criterion for the selection of meta-analysis included studies. Case reports, editorials, commentaries, reviews, studies with insufficient data, and non-English articles were excluded. All steps were double checked and in case of disagreement, a third reviewer opinion was obtained.

### Data Extraction and Quality Assessment

Two reviewers (AS and MH) designed an agreed data extraction form and extracted the data of all eligible studies. The first author’s name, date of publication, type of study, country of study conduction, mean age, study population, type of surgery, site of surgery, the reported or investigated Clavien-Dindo classification were extracted.

Type of surgeries based on urgency: According to the NCEPOD association, the surgeries can be categorized into four levels of urgency (18):

a. Immediate: Surgeries must perform in life-threatening conditions within minutes after the diagnosis.
b. Urgent: Surgeries in potentially life-threatening conditions which must perform within hours after the diagnosis.
c. Expediated: Surgeries which should perform early but it can be delayed for days.
d. Elective: Surgeries which are planned to perform at the optimum condition of the patient.

Clavien-Dindo classification of post operative complications: Surgical complications are categorized into five grades due to Clavien-Dindo classification (19):

I. Grade 1 is defined as any deviation from the normal expected recovery path, which does not require any specific treatment; the analgesics, anti-emetics, electrolyte, diuretics, physiotherapy, and the bedside wound infections opening are the allowed prescription and interventions.
II. Grade 2 is any medical treatment but the above.
III. Grade 3 is any intervention; Including surgical, radiologic and endoscopic.
IV. Grade 4 is any life-threatening complication, required intensive care.
V. Grade 5 is death of the patient.

### Quality Assessment

Two other reviewer (FS and YF) blindly assessed the quality of the included articles by the ROBINS-I (“Risk Of Bias In Non-randomized Studies - of Interventions”) (20). The study population, methods of matching and controlling the confounding factors, the complication reporting system, follow up period, and statistical analysis were assessed with great concern.

### Data Synthesis and Analysis

Statistical analyses were performed with Comprehensive Meta-Analysis software, version v3.7z (Biostat Inc, Englewood, USA). The heterogeneity was assessed by Cochran’s Q and the I^2^ statistic. According to the estimated heterogeneity of the true effect sizes for each grade of complications, either the random effect model or the fixed effect model was used. The odds of complications between pre-COVID and COVID era groups with 95% confidence intervals (CI) were calculated. Begg’s and Egger’s tests were used to assess the publication bias and funnel plots illustrated it (*p* < 0.05 was indicative for statistically significant publication bias).

## Result

The primary search result in databases revealed 909 articles, which finally 34 articles were included for qualitative analysis which 11 out of them were eligible for quantitative analysis (Figure 1). A summary of all included articles is provided in Table 1. A total of 19,137 patients (3522 patients in pre COVID era and 15615 patients during COVID era) were assessed in meta-analysis. According to ROBINS-I tool, the included studies for both systematic review and meta-analysis had low risk of bias (Supp.1). There was unsignificant publication bias which is expressed as Begg’s and Egger’s indexes (Table 2). Also funnel plots are provided in supp.2 to illustrate the publication bias.

**Table 1.** Characteristics of the included studies. The first 11 studies are used for meta-analysis. med: Median; NM: Not mentioned.

| First Author | Date of publication | Country | Type of study | Population | Pre COVID Vs. COVID Era | Male: Female ratio | Mean age (years) |
| --- | --- | --- | --- | --- | --- | --- | --- |
| Fonseca et al. (21) | 2020, Nov | Brazil | Cohort | 118 | 82: 36 | 47: 71 | 35.5 |
| Tartaglia et al. (22) | 2020, Dec | USA | Cohort | 143 | 91: 52 | 78: 65 | 64.85 |
| Würnschimmel et al. (23) | 2020, Aug | Germany | Cohort | 784 | 447: 337 | 784: 0 | 64 |
| Sartori et al. (24) | 2021, July | Italy | Cohort | 1336 | 791: 546 | 449: 388 | 37.84 |
| Jain et al. (25) | 2021, Jan | India | Cohort | 127 | 59: 68 | 84: 44 | 52.4 |
| Borgstein et al. (26) | 2021, Apr | Netherlands | Cohort | 307 | 168: 139 | 257: 50 | 66.45 |
| Hugo et al.(27) | 2021, Aug | Switzerland | Cohort | 214 | 115: 99 | 111: 103 | 42.5 |
| Malik et al. (28) | 2021, Jun | UK | Cohort | 71 | 32: 39 | 43: 28 | 66.8 |
| Rashid et al. (29) | 2021, Aug | UK | Cohort | 32 | 10: 22 | 16: 6 | 74.2 |
| Panda et al. (30)<sup>1</sup> | 2021, Sep | India | Cohort | 64 | 32: 32 | 52: 12 | 47.34 |
| Fernández-Martínez et al. (31) | 2021, Jun | Spain | Cohort | 235 | 135: 100 | NM | NM |
| Vusirikala et al. (32) | 2021, Jul | UK | Cohort | 104 | 0: 104 | 59: 45 | 59 |
| Iqbal et al. (33) | 2020, Oct | UK | Cohort | 153 | 0: 153 | 58: 95 | 57 |
| Rajasekaran et al. (34) | 2020, Sep | UK | Cohort | 56 | 0: 56 | 30: 26 | 57 (med) |
| Wahed et al. (35) | 2020, Aug | UK | Cohort | 19 | 0: 19 | 15: 4 | 70 (med) |
| Minervini et al. (36) | 2021, Apr | Italy | Cohort | 1943 | 0: 1943 | NM | 67 (med) |
| Romics et al. (37) | 2020, Nov | UK | Cohort | 179 | 0: 179 | 0: 179 | 54 (med) |
| Singhal et al. (38) | 2021, May | UK | Cohort | 7704 | 0: 7704 | 1886: 519 | 40.35 |
| Clifford et al. (39) | 2021, Jun | UK | Cohort | 21 | 0: 21 | 3: 18 | 53 (med) |
| Dinçer et al. (40) | 2021, Sep | Turkey | Cohort | 223 | 0: 223 | 112: 111 | 48.5 |
| Jeannon et al. (41) | 2021, Jan | UK | Cohort | 69 | 0: 69 | NM | 52 |
| Kulle et al. (42) | 2021, Feb | Turkey | Cohort | 404 | 0: 404 | NM | 61 (med) |
| Kasivisvanathan et al. (43) | 2020, Oct | UK | Cohort | 500 | 0: 500 | 327: 173 | 62.5 (med) |
| ROMANZI et al.(44) | 2020, Oct | Switzerland | Cohort | 18 | 0: 18 | 8: 10 | 73.2 |
| Romanzi et al. (45) | 2021, Feb | Italy | Cohort | 13 | 0: 13 | 5: 8 | 80 |
| Zagra et al. (46) | 2021, Jun | Italy | Cohort | 38 | 0: 38 | 12: 26 | 81 |
| Monroy-Iglesias et al. (47) | 2021, Mar | UK | Cohort | 3037 | 1560: 1477 | 1707: 1330 | 62.49 |
| Brar et al. (48) | 2021, Jul | UK | Cohort | 47 | 0: 47 | 31: 16 | NM |
| Gammeri et al. (49) | 2020, Jul | UK | Cohort | 309 | 0: 309 | 142: 167 | 61.9 |
| Castellvi et al. (50) | 2020, Aug | Spain | Cohort | 171 | 0: 171 | 97: 83<sup>2</sup> | 65.7 |
| Rottoli et al. (51) | 2021, Jun | Italy | Cohort | 91 | 0: 91 | 59: 32 | 43 |
| Das et al. (52) | 2021, Mar | India | Cohort | 457 | 0: 457 | NM | 50 (med) |
| Evans et al. (53) | 2021, Jul | UK | Cohort | 23 | 0: 23 | 16: 7 | 65 |
| Santiago et al. (54) | 2020, Jul | Spain | Cohort | 126 | 0: 126 | NM | 67 (med) |

<sup>1</sup> The matched groups are considered.
<sup>2</sup> Some of the patients managed non- operatively. The study population refers to the number of surgery but the gender specification has considered all the patients.

**Table 2.**
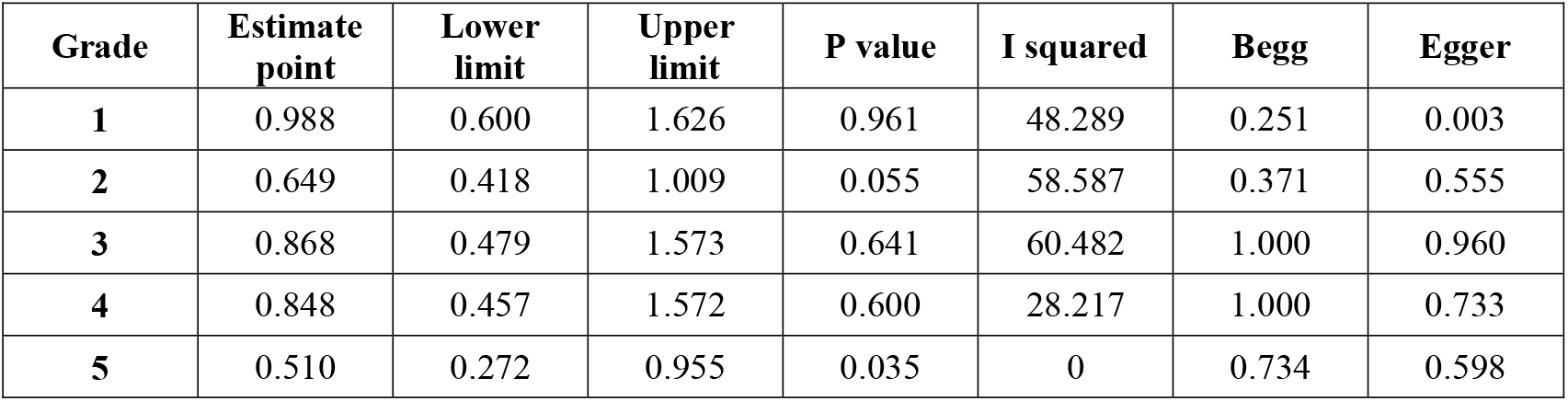
The Meta- analysis of the incidence of each grade of surgical complications and its publication bias due to Clavien-Dindo classification.

| Grade | Estimate point | Lower limit | Upper limit | P value | I squared | Begg | Egger |
| --- | --- | --- | --- | --- | --- | --- | --- |
| 1 | 0.988 | 0.600 | 1.626 | 0.961 | 48.289 | 0.251 | 0.003 |
| 2 | 0.649 | 0.418 | 1.009 | 0.055 | 58.587 | 0.371 | 0.555 |
| 3 | 0.868 | 0.479 | 1.573 | 0.641 | 60.482 | 1.000 | 0.960 |
| 4 | 0.848 | 0.457 | 1.572 | 0.600 | 28.217 | 1.000 | 0.733 |
| 5 | 0.510 | 0.272 | 0.955 | 0.035 | 0 | 0.734 | 0.598 |

**Figure 1.**
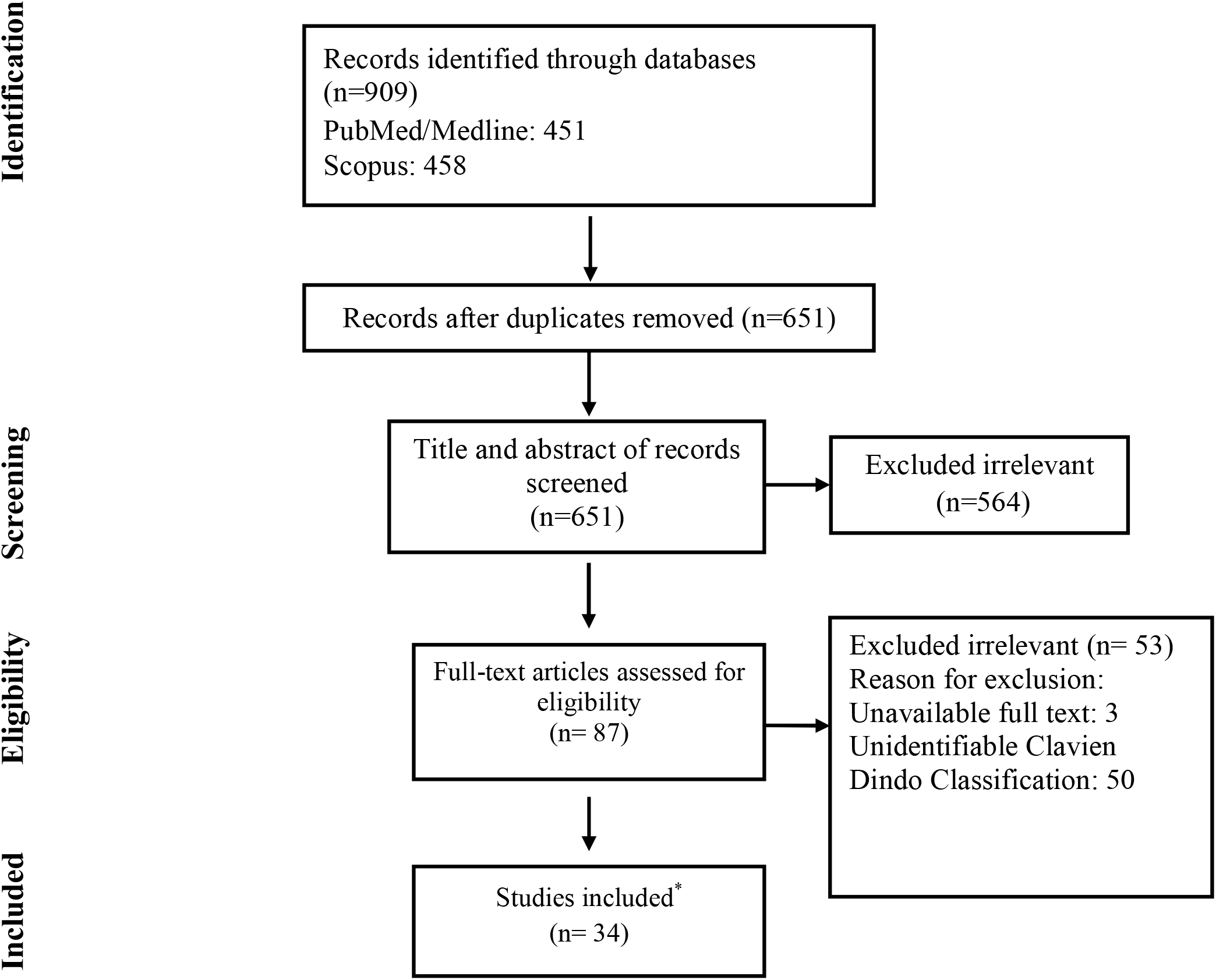
Flow chart of study selection for inclusion in the systematic review *Total number of 34 articles reviewed qualitatively and included in systematic review, 11 of them were assessed quantitively for meta-analysis too.

### The urgency of operations and related organ systems

Elective surgeries (including cancer surgeries) were the most common type of applied surgeries (86.32%), followed by urgent (11.22%), emergent and expediated surgeries.

Non-vascular abdominal surgeries, urologic/ gynecologic surgeries, and the cardiothoracic surgeries were the most common reported surgeries. All reported surgeries based on the related organ-system is noted in Table 3.

**Table 3.** The performed surgeries by urgency and related organ- system

| Categorization basis | Categories | Absolute frequency (n) | Relative frequency (%) |
| --- | --- | --- | --- |
| Urgency | Immediate | 327 | 1.71 |
|  | Urgent | 2150 | 11.22 |
|  | Expedited | 145 | 0.76 |
|  | Elective | 16547 | 86.32 |
| Field of surgery | Neurosurgery | 52 | 0.28 |
|  | Cardiothoracic | 1321 | 7.19 |
|  | Non-vascular Abdominal | 9594 | 52.19 |
|  | Gynaecology / Urology | 5180 | 28.18 |
|  | Breast | 649 | 3.53 |
|  | Orthopaedics | 260 | 1.41 |
|  | Head and neck | 819 | 4.46 |
|  | Plastic | 185 | 1.01 |
|  | Ophthalmic | 4 | 0.02 |
|  | Others* | 319 | 1.74 |

\*Others includes soft tissue and bone sarcoma surgery, endocrine surgery, and other unspecified surgeries. The discrepancy between the total number of surgeries between two categorization system is due to unnoted surgical fields in some of the reviewed studies.

### Post operative complications

Post operative complications are reported based on Clavien-Dindo classification. Table 4. Demonstrates the incidence of post operative complications in reviewed articles during the COVID era (Table 4). The incidence of each grade of post operative complication based on Clavien-Dindo classification, has been compared and meta-analyzed between pre COVID era and COVID era, illustrated in figures 2-6. There was no significant difference between any grades of post operative complications between pre COVID era and COVID era (Table 2, 4, 5, Figures 2-6).

**Table 4.** The incidence of post operative surgical complications due to Clavien- Dindo classification during the COVID era.

| First author | COVID Era sample size | Clavien- Dindo Classification grades |  |  |  |  |
| --- | --- | --- | --- | --- | --- | --- |
|  |  | 1 | 2 | 3 | 4 | 5 |
| Fonseca et al. (21) | 36 | 5 | 3 | 3 | 0 | 0 |
| Tartaglia et al. (22) | 52 | 4 | 7 | 1 | 5 | 12 |
| Würnschimmel et al. (23) | 337 | 21 | 26 | 13 | 2 | 0 |
| Sartori et al. (24) | 546 | 402 | 108 | 29 | 3 | 1 |
| Jain et al. (25) | 68 | 1 | 0 | 5 | 1 | 0 |
| Borgstein et al. (26) | 139 | 5 | 33 | 25 | 25 | 5 |
| Hugo et al.(27) | 99 | 4 | 5 | 3 | 0 | 0 |
| Malik et al. (28) | 39 | 0 | 7 | 4 | 0 | 0 |
| Rashid et al. (29) | 22 | 18 | 4 | 0 | 0 | 0 |
| Panda et al. (30)<sup>3</sup> | 32 | 2 | 10 | 2 | 1 | 0 |
| Fernández□Martínez et al. (31) | 100 | 0 | 9 | 10 | 9 | 7 |
| Vusirikala et al. (32) | 104 | 5 | 5 | 1 | 0 | 0 |
| Iqbal et al. (33) | 153 | 1 | 6 | 0 | 0 | 0 |
| Rajasekaran et al. (34) | 56 | 2 | 0 | 11 | 0 | 0 |
| Wahed et al. (35) | 19 | 0 | 5 | 1 | 1 | 0 |
| Minervini et al. (36) | 1943 | 26 | 32 | 18 | 0 | 1 |
| Romics et al. (37) | 179 | 7 | 2 | 4 | 0 | 0 |
| Singhal et al. (38) | 7704 | 193 | 158 | 160 | 46 | 10 |
| Clifford et al. (39) | 21 | 0 | 2 | 0 | 0 | 0 |
| Dinçer et al. (40) | 223 | 23 | 35 | 36 | 0 | 8 |
| Jeannon et al. (41) | 69 | 4 | 7 | 0 | 0 | 0 |
| Kulle et al. (42) | 404 | 11 | 29 | 32 | 2 | 5 |
| Kasivisvanathan et al. (43) | 500 | NM | NM | 16 | 14 | 3 |
| ROMANZI et al.(44) | 18 | 0 | 1 | 1 | 0 | 0 |
| Romanzi et al. (45) | 13 | 0 | 1 | 0 | 0 | 0 |
| Zagra et al. (46) | 38 | 3 | 6 | 16 | 6 | 5 |
| Monroy-Iglesias et al. (47) | 1477 | 92 | 81 | 37 | 26 | 3 |
| Brar et al. (48) | 47 | 2 | 5 | 1 | 0 | 0 |
| Gammeri et al. (49) | 309 | 13 | 12 | 2 | 0 | 0 |
| Castellvi et al. (50) | 171 | 26 | 0 | 4 | 0 | 0 |
| Rottoli et al. (51) | 91 | 1 | 13 | 5 | 1 | 0 |
| Das et al. (52) | 457 | 375 | 78 | 2 | 0 | 2 |

<sup>3</sup> The matched groups are considered.

|  |  |  |  |  |  |  |
| --- | --- | --- | --- | --- | --- | --- |
| Evans et al. (53) | 23 | 6 | 4 | 0 | 0 | 0 |
| Santiago et al. (54) | 126 | 9 | 4 | 1 | 1 | 0 |

**Table 5.**
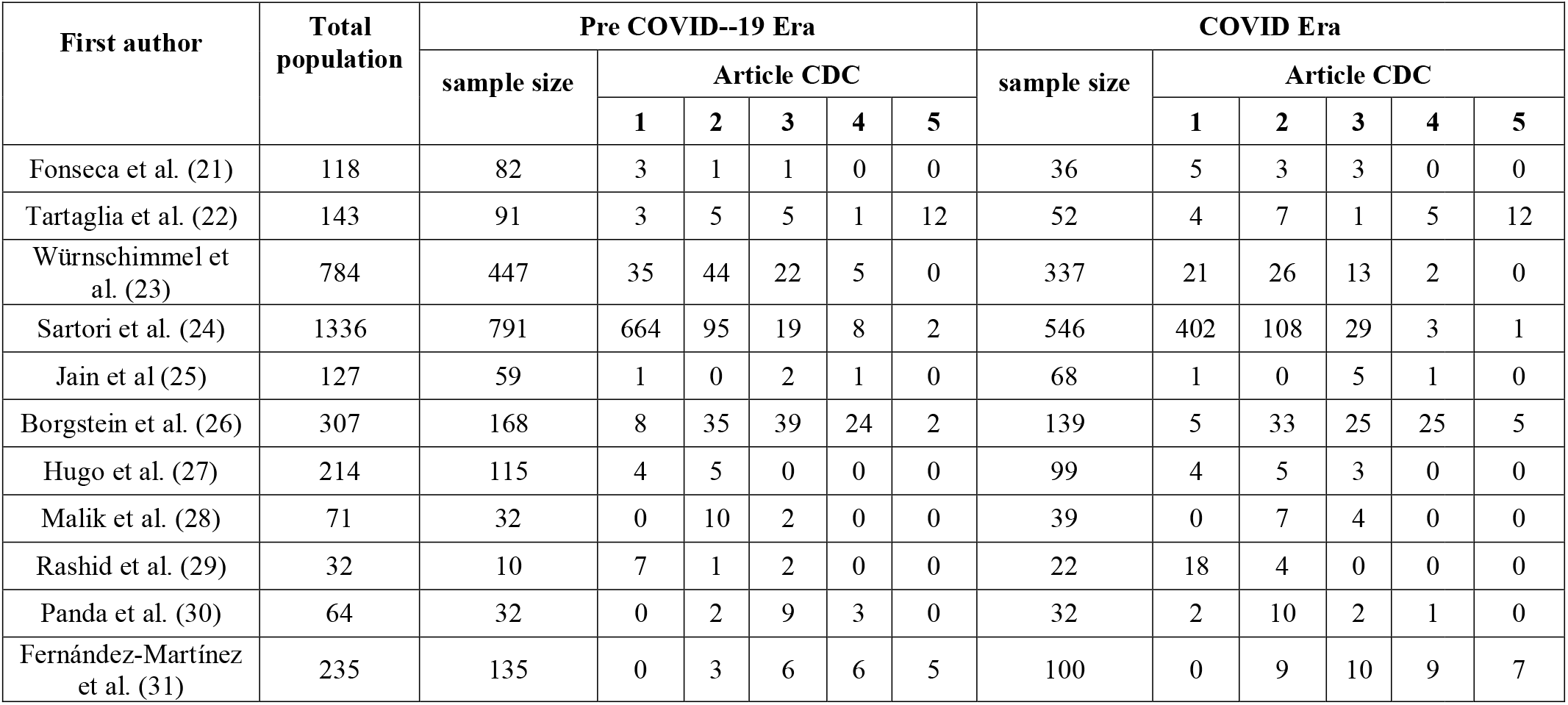
The incidence of surgical complications due to Clavien- Dindo classification before and during the COVID -19 era.

| First author | Total population | Pre COVID--19 Era |  |  |  |  |  | COVID Era |  |  |  |  |  |
| --- | --- | --- | --- | --- | --- | --- | --- | --- | --- | --- | --- | --- | --- |
|  |  | sample size | Article CDC |  |  |  |  | sample size | Article CDC |  |  |  |  |
|  |  |  | 1 | 2 | 3 | 4 | 5 |  | 1 | 2 | 3 | 4 | 5 |
| Fonseca et al. (21) | 118 | 82 | 3 | 1 | 1 | 0 | 0 | 36 | 5 | 3 | 3 | 0 | 0 |
| Tartaglia et al. (22) | 143 | 91 | 3 | 5 | 5 | 1 | 12 | 52 | 4 | 7 | 1 | 5 | 12 |
| Würnschimmel et al. (23) | 784 | 447 | 35 | 44 | 22 | 5 | 0 | 337 | 21 | 26 | 13 | 2 | 0 |
| Sartori et al. (24) | 1336 | 791 | 664 | 95 | 19 | 8 | 2 | 546 | 402 | 108 | 29 | 3 | 1 |
| Jain et al (25) | 127 | 59 | 1 | 0 | 2 | 1 | 0 | 68 | 1 | 0 | 5 | 1 | 0 |
| Borgstein et al. (26) | 307 | 168 | 8 | 35 | 39 | 24 | 2 | 139 | 5 | 33 | 25 | 25 | 5 |
| Hugo et al. (27) | 214 | 115 | 4 | 5 | 0 | 0 | 0 | 99 | 4 | 5 | 3 | 0 | 0 |
| Malik et al. (28) | 71 | 32 | 0 | 10 | 2 | 0 | 0 | 39 | 0 | 7 | 4 | 0 | 0 |
| Rashid et al. (29) | 32 | 10 | 7 | 1 | 2 | 0 | 0 | 22 | 18 | 4 | 0 | 0 | 0 |
| Panda et al. (30) | 64 | 32 | 0 | 2 | 9 | 3 | 0 | 32 | 2 | 10 | 2 | 1 | 0 |
| Fernández-Martínez et al. (31) | 235 | 135 | 0 | 3 | 6 | 6 | 5 | 100 | 0 | 9 | 10 | 9 | 7 |

**Figures 2- 6.**
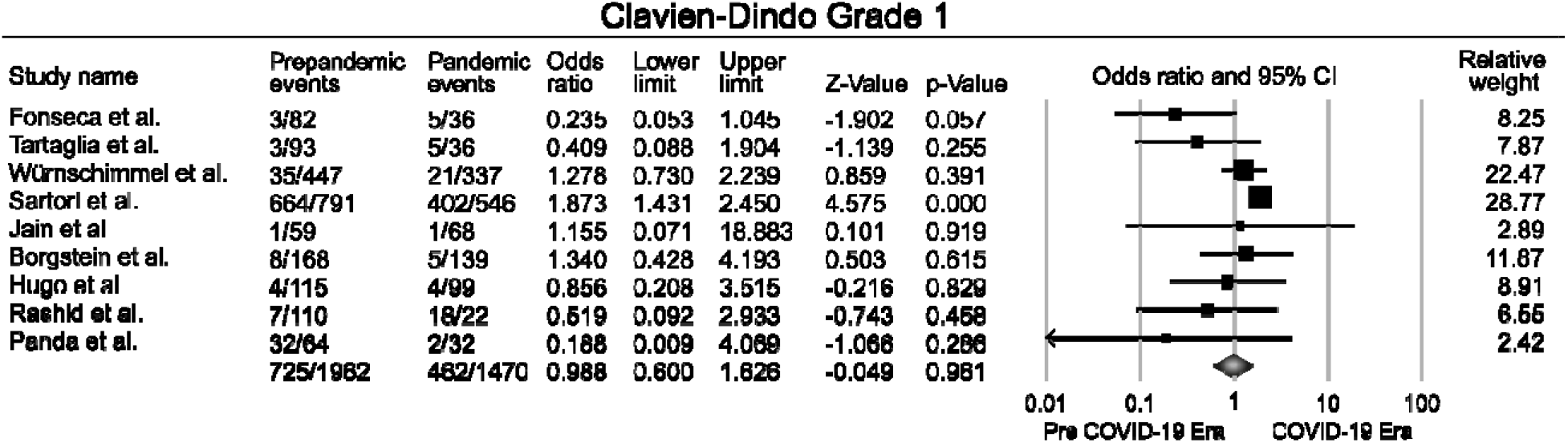
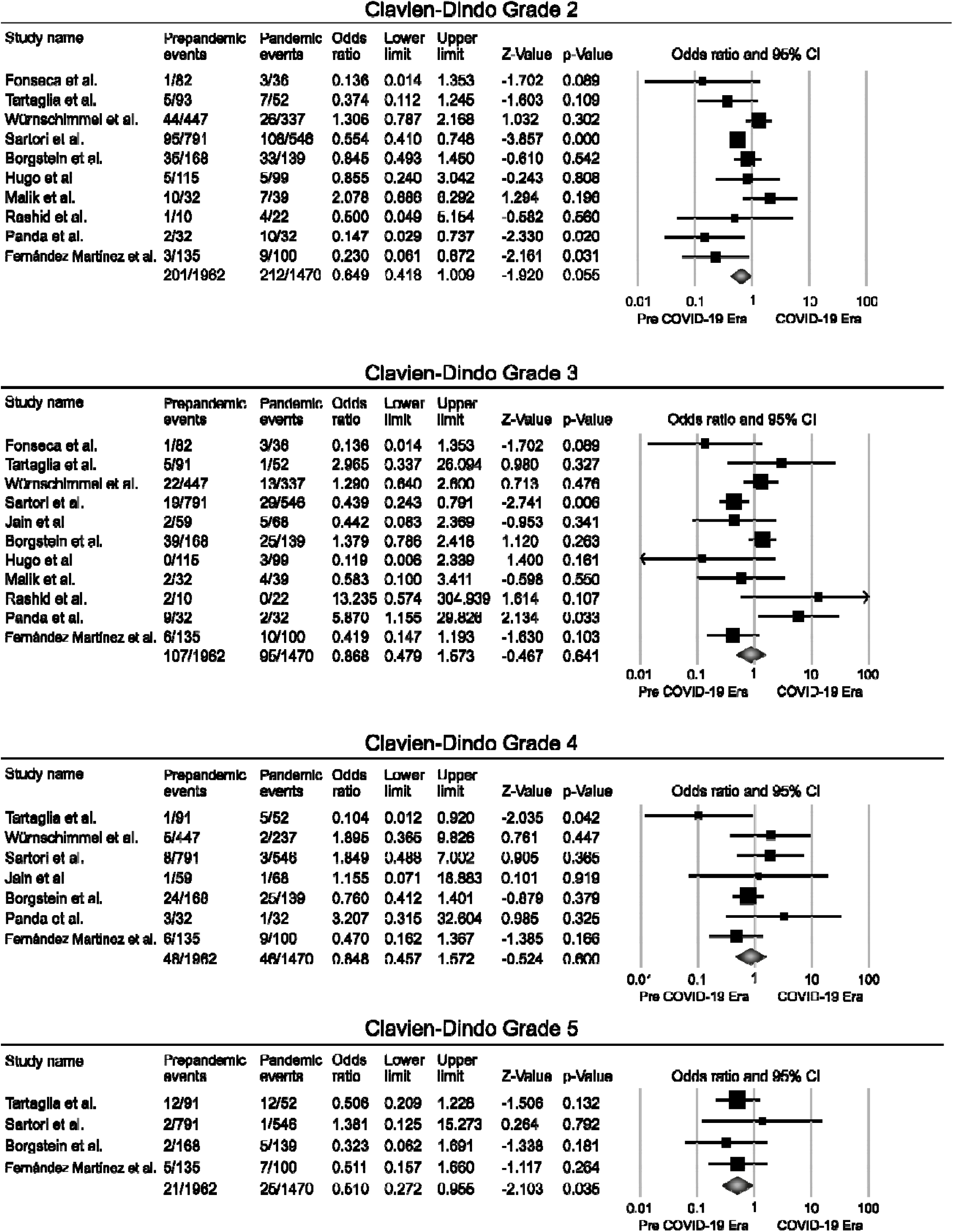
The Forest Plot of each grade.

**Figure 7.**
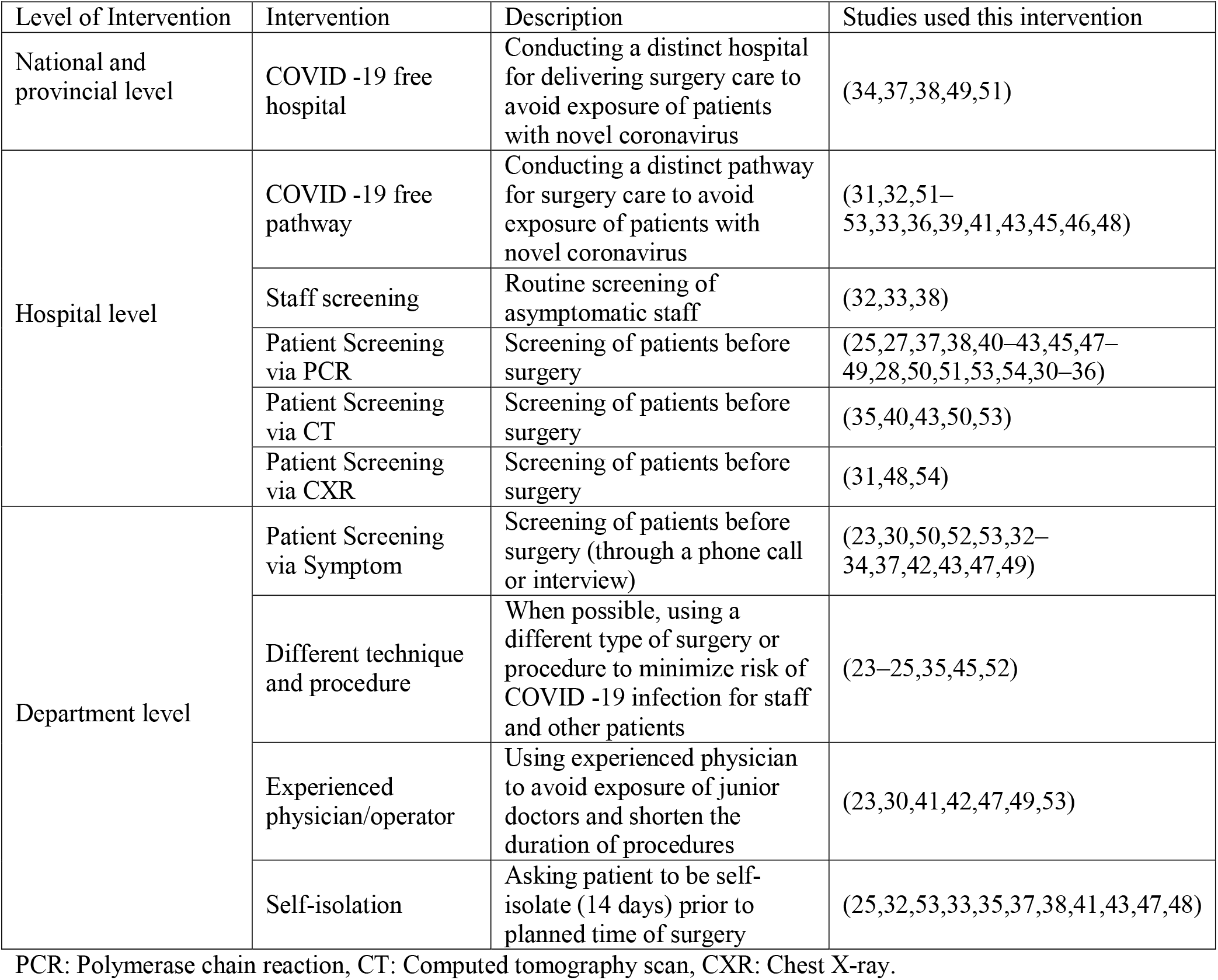
Adopted strategies and interventions during the pandemic to continue delivering surgery care PCR: Polymerase chain reaction, CT: Computed tomography scan, CXR: Chest X-ray.

## Discussion

Since the COVID -19 outbreak in early 2020, almost all healthcare facilities worldwide have faced devastating damages from this pandemic. It has altered proper medical care by delays in diagnosis, treatment, and follow-up of patients(35). Massive employment of health care resources resulted in canceling and postponing surgeries. Some studies reported an increase in postoperative complications(24,55,56). As far as we aware this study is among first systematic reviews studying on complication of surgeries during the pandemic so far. we collected and reviewed data to compare the rate of postoperative complications as classified by Clavien-Dindo Classification (CDC) before and during the COVID -19 outbreak. Although the surgery complications were comparable (grades 1 to 4), the post-operative mortality (grade 5) was higher during the pandemic compared to before the outbreak. The more advanced stage of the disease, delay in treatment, and more complicated patient selection may explain the lower survival after the surgery.

Several studies dedicate higher post operative morbidity and mortality among SARS-CoV-2 positive patients (56–59); but the impact of COVID -19 on surgical practice is not limited to SARS-CoV-2 positive patients. According to our meta-analysis, the COVID -19 negative patients who underwent surgery during the COVID -19 era are also endangered of greater mortality (60). Delay in hospital referral due to public fear of COVID -19, has led to more critical condition of patients at the admission time which was accompanied by more complicated surgical state and higher mortality, especially for emergency surgeries. (24,27,61,62). On the other hand, health care providers preferred close observation and conservative management to surgery in patients with younger age and better overall health condition (31,63,64) So the majority of operations were performed on elder adults with multiple comorbidities and more critical conditions with poorer prognosis (24,33,34,46,52,65).

Studies from different contexts were included in our review. Different centers, countries, timelines, types of surgeries and guidelines were observed, which may affect the surgical complications. Changes in resources availability were also presented in most studies (REF?). Three out of the ten studies in our meta-analysis suggested significantly higher rate of post-operative complications (22) and one of the two reported more severe complications in the COVID -19 era compared to the control cohort (24). Both studies were conducted in the first COVID -19 wave in Italy which was one of the first countries to experience the SARS-COV-2 outbreak. Above all, these studies did not claim any specific screening or preventing protocols for COVID -19. In contrast, with the use of protection protocols, Panda et al. found a significant decrease in major post-operative complications during pandemic, and in a matched pair analysis, lower rate of complications (30). Evidence suggests that surgery care can be continued with adopting preventive strategies and protocols, even in the COVID -19 peak (23,26–29,42,49,54).

As **Figure 8** depicts, many preventive strategies were implemented in an attempt to continue routine surgery care. A study on 7704 bariatric patients from 42 countries admitted in 185 centers showed 50% and 80% of centers practiced self-isolation and screening before surgery, respectively. Half of centers provided FFP3/N95 masks in the operation room and staff-screening was performed in third of hospitals. They found same complication rate during the pandemic and concluded it is safe to continue surgery care using preventive protocols (38).

Moreover, COVID Surg Collaborative study on 9171 patients from 447 hospitals proved lower respiratory complication in hospitals with COVID -19-free pathway (66). A variety of protocols is evident in the literature at national, hospital, and department levels (**Figure 8**). Although some screening and protective measures are considered to be more effective, some of them are not feasible in low-resourced contexts. However, proper decisions at department and hospital level could successfully prevent complications regardless of shortage of resources.

Approaches of treatment in different settings were also affected by the pandemic and resulted in new guidelines and recommendations (64). Based on previous experience from viruses such as HPV, some initial guidelines had recommended limitation of laparoscopy procedures as it may carry the risk of surgical staff contamination (67). However, with employment of proper ultrafiltration and protection methods, studies not following this recommendation did not observe such side effect (29,54) Moreover, recent guidelines recommended conservative treatment of uncomplicated appendicitis as a safe option with low failure rate especially in the COVID -19 pandemic (68,69). Prioritization of the patients, ICU preserving techniques, laparotomy with regional instead of general anesthesia (42,45) more extensive use of robotic surgery (70), and distant/home patient monitoring and follow-up were put into test. Despite all limitations, it seems that the means and measures for lowering post-operative complications are effective. m-RNA vaccines have been created for COVID -19 offering hope for HIV prevention (71). Likewise, we hope development and employment of these new strategies in healthcare and surgery could lead to positive effect not only during pandemics but also in the future practice.

Studies in the available literature confirmed a decrease in admission rates (21,22,24,32) which may be a result of patients’ reluctancy to admission in fear of COVID -19. Prognostic factors and corresponding indices such as complications at presentation (e.g., perforation, abscess, etc.), Mannheim Peritonitis Index, American Association for the Surgery of Trauma (ASST) scores, and pathologic findings were significantly in favor of complexity (21,22,24,27,28,42) and severity (26,42) of the diseases in the COVID era. Changes in rate of admissions can also cause outcome bias. On one hand, elective surgeries (which have better prognosis) significantly decreased in some contexts (22,29,32). On the other hand, less surgical admission along with strict COVID -19 protocols resulted in less operations and slower turnover of the patients which can be the reason for the favorable outcomes.

Apart from the potential risk of COVID -19 itself, the recent pandemic has posed unprecedented challenges in terms of finance, resources, disease severity, and outcomes for the healthcare system around the world. Cancelation and subsequent long waiting list of elective surgeries, low capacity of hospital and ICU beds especially in the first months of pandemic can negatively affect surgeries (12,13). The importance in preventing unnecessary delays in non-elective and cancer surgeries is irrefutable (72). Thus, preserving enough capacity for cancer surgery along with ongoing emergent surgeries is of great concern. In a center with no preoperative screening in asymptomatic patients, extraordinary hospital and ICU bed capacity along with protocols resulted in favorable outcomes (23). Overall delay in treatment may put the patient’s life at risk and may explain some extent of higher mortality during the pandemic.

There are limitations when interpreting the results of this study. Many of included studies was conducted in developed countries, whereas studies from developing countries are scarce and no study was available on African population. This is important since the under-resourced countries face shortage of screening tools, lack of resource to change infrastructures, and less flexibility to adapt new strategies. Moreover, small samples were available for some of surgery fields such as ophthalmology, therefore the implementation of the results should be done with caution.

In conclusion, although delivering routine surgery was challenging, the perioperative complications during the pandemic remained identical to the pre-pandemic era. Precise patient selection and adopting preventive strategies helped deliver surgeries during the outbreak of COVID -19 while limiting the capacity of operations and admissions. Studies from middle- and low-income countries are scarce, where implementing new protocols is challenging. Future studies are warranted to confirm these results.

## Supporting information

Supplemental figures, Funnel plots

## Data Availability

All data produced in the present study are available upon reasonable request to the authors.

## Author Contribution

Guarantor of article: YF

study concept and study supervision: MN study design: MN, YF

manuscript drafting: FS, AS, MH, BM

manuscript edition and final approval: all authors

data analysis and visualization: YF, AS

data collection and/or data interpretation: all authors

