## Supplementary figures and images for "The comparison of Post-Operative Complications pre Covid era versus during Covid-Era based on Clavien-Dindo-classification: A Systematic Review and Meta-Analysis"

### Supplemental figures, Funnel plots

**Supp 2.** Publication bias of articles reporting each grade, illustrated as funnel plots.

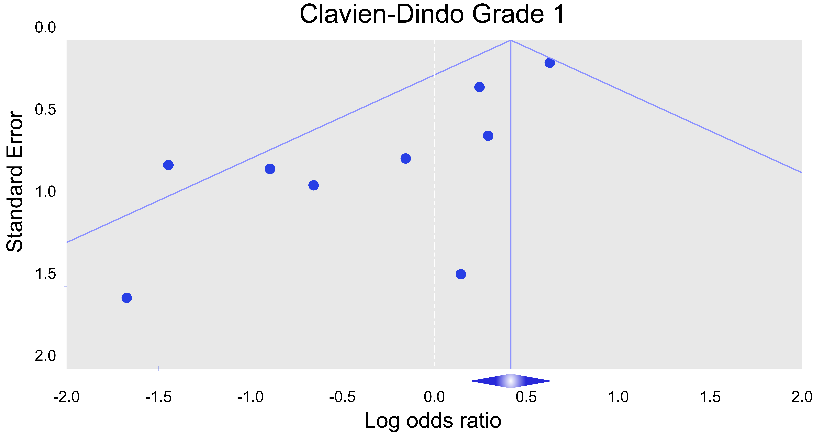

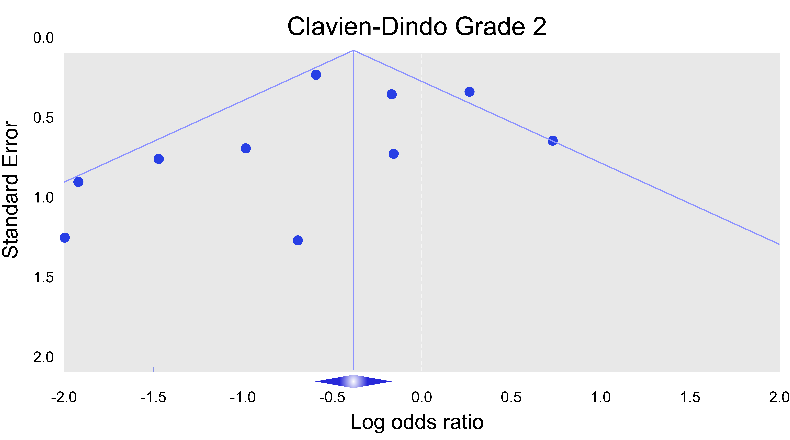

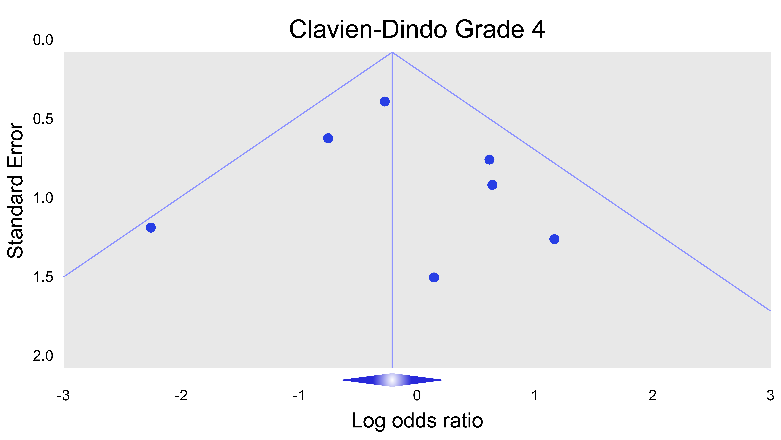

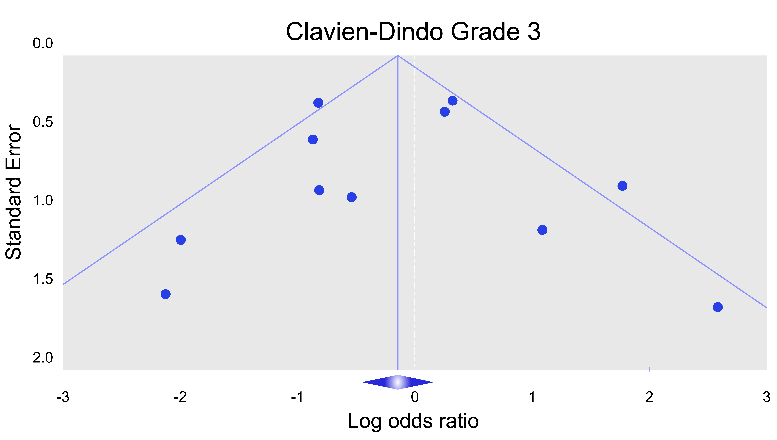

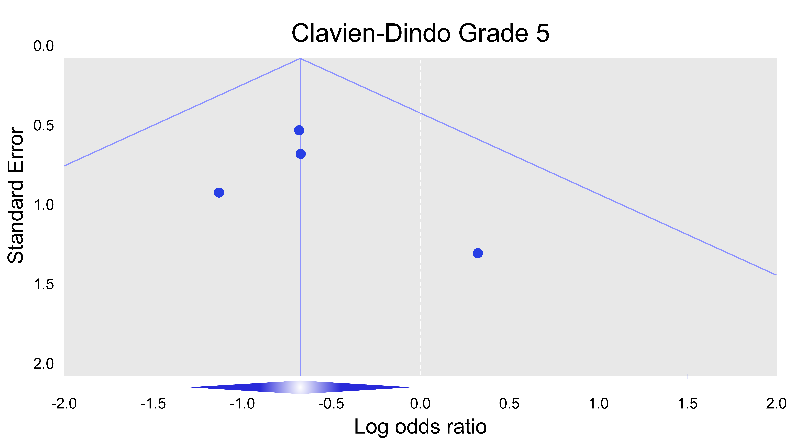
